# Cohort-scale automated patch clamp data improves variant classification and penetrance stratification for *SCN5A*-Brugada Syndrome

**DOI:** 10.1101/2025.03.09.25323605

**Authors:** Matthew J. O’Neill, Joanne G. Ma, Jessa L. Aldridge, Joseph F. Solus, Genevieve R. Harvey, Paige H. Roberson, Julian Barc, Connie R. Bezzina, Dan M. Roden, Roddy Walsh, Jamie I. Vandenberg, Andrew M. Glazer, Chai-Ann Ng

## Abstract

**Background:** Brugada Syndrome (BrS) is an inherited arrhythmia disorder that causes an elevated risk of sudden cardiac death. Approximately 20% of patients with BrS have rare variants in *SCN5A*, which encodes the cardiac sodium channel Na_V_1.5. Genetic workup of BrS is often complicated by *SCN5A* variants of uncertain significance (VUS) and/or incomplete penetrance.

**Methods:** We analyzed all 252 missense and in-frame insertion/deletion *SCN5A* variants from a previously published large cohort of BrS cases (n=3,335 patients) using a calibrated high-throughput automated patch clamp (APC) assay. Variant functional *Z*-scores were assigned evidence levels ranging from BS3_moderate (normal function) to PS3_strong (loss-of-function), as defined by American College of Medical Genetics and Genomics criteria. Functional evidence was combined with population frequency, hot-spot, case counts, protein length changes, and *in silico* predictions. Odds ratios of BrS case-control enrichment and penetrance for BrS were calculated from variant frequencies in the BrS cohort and in gnomAD.

**Results:** Most variants (146/252) were functionally abnormal (*Z* ≤ -2), with 100 having severe loss-of-function (*Z* ≤ -4). Functional evidence enabled the reclassification of 110 of 225 VUS; 104 to likely pathogenic and 6 to likely benign. *SCN5A* variants with loss-of-function were mainly localized to the transmembrane domains, especially the regions comprising the central pore. *SCN5A* variant penetrance was proportional to the severity of loss-of-function; variants with Z ≤ -6 had penetrance of 24.5% (15.9 – 37.7% CI) and an odds ratio of 501 for BrS.

**Conclusions:** This cohort-scale APC dataset stratifies *SCN5A* variants found in BrS patients into normal function “bystander” variants that have a low risk for BrS and loss-of-function variants that have a high risk for BrS. Functional data can be integrated with other criteria to reclassify a substantial fraction of VUS. The dataset helps clarify the *SCN5A*-BrS relationship and will improve the diagnosis and clinical management of BrS probands and their families.

## Introduction

Brugada Syndrome (BrS; MIM:601144) is a heritable arrhythmia syndrome characterized by ST-segment elevation in the right precordial leads on the electrocardiogram and increased risk for severe ventricular arrhythmias and sudden death.^1^ Early diagnosis of BrS is critical, as arrhythmias or sudden cardiac death can be the first manifestation of the disease.^2, 3^ For individuals with BrS, treatments such as medication, ablations, or an implanted cardioverter defibrillator can be lifesaving.^4^ Although large-effect size variants in over 20 genes have been suggested to cause BrS, a ClinGen expert panel found that rare variants in only *SCN5A* have definitive evidence for causing BrS.^5^ *SCN5A* encodes the cardiac voltage-gated sodium channel Na_V_1.5, which is responsible for most of the depolarization at the start of the cardiac action potential.^6^ Rare loss-of-function (LOF) variants in *SCN5A* are present in 15-30% of BrS cases, depending on ancestry and disease sensitivity.^7^ A high common genetic variant burden can also contribute to BrS risk or modify the penetrance of rare *SCN5A* variants.^1, 8, 9^ Notably, BrS has a large degree of incomplete penetrance and only a fraction of carriers with the same rare genetic variant present with BrS.^8^

The American College of Medical Genetics and Genomics/Association for Molecular Pathology (ACMG/AMP) framework established criteria for variant classification in medically important genes, including *SCN5A*.^10, 11^ These criteria include allele frequency (AF), co-segregation with disease, location in protein hot-spots, *in silico* prediction tools, and laboratory functional experiments.^10^ Classifications range from Benign/Likely Benign (B/LB) to Pathogenic/Likely Pathogenic (P/LP). However, most missense variants in *SCN5A* cannot be definitively classified and are instead labelled Variants of Uncertain Significance (VUS).

Recently, the genetic characteristics of a cohort of 3,335 BrS patients has been published.^9, 12^ The cohort includes 614 individuals with rare heterozygous *SCN5A* variants, spanning 353 unique variants—103 predicted LOF variants (*e.g.* nonsense, frameshift, or splice-site variants) and 252 missense or in-frame insertion/deletion (indel) variants.^12^ This large patient dataset provides an opportunity to improve variant classifications and estimate variant-specific BrS penetrance. Several improved classification approaches have been proposed. A modified hot-spot criteria (PM1) based on each variant’s relative frequency in cases and controls which has helped to reduce the number of VUS;^12^ however, there are functionally normal variants within each hot-spot.^13^ In addition, *in silico* predictions such as AlphaMissense^14^ and REVEL^15^ were calibrated by ClinGen as providing up to a strong level of evidence strength for variant classification (PP3/BS4).^16^ Variant *in vitro* functional data (PS3/BS3) can also improve variant classifications.^17^

Ideally, cohort datasets would be paired with comprehensive *in vitro* functional datasets to support variant interpretation. However, in practice, functional data are only sparsely available for inherited arrhythmia syndromes, where traditional manual patch clamp studies of ion channel variants are time-intensive and low-throughput. To overcome this limitation, we developed a high-throughput automated patch clamp (APC) assay to investigate *SCN5A*-BrS variant function.^18–20^ We recently calibrated the assay at two independent sites using a large set of benign and BrS pathogenic/likely pathogenic variant controls,^19^ following guidelines recommended by the ClinGen Sequence Variant Interpretation working group.^21^ The assay accurately distinguished benign and pathogenic variants, supporting the application of a strong level of evidence for LOF (PS3) or moderate evidence for normal-function variants (BS3_moderate).

Here, we deployed the *SCN5A*-BrS functional assay at cohort-scale to facilitate the implementation of genetic and precision medicine. We provided calibrated APC data as *Z-* scores for the entire set of 252 rare *SCN5A* missense and in-frame indel variants previously identified in the published cohort of 3,335 BrS patients.^12^ We then determined the value of calibrated *Z*-scores as functional evidence to reclassify *SCN5A*-BrS cohort variants in conjunction with other criteria including population frequency, hot-spot, case-enrichment, protein-length, and *in silico* tools. Lastly, we calculated estimates of the relationship between variant-specific function and both case-control enrichment and BrS penetrance using the cohort of 3,335 BrS patients.

## Methods

### High-throughput Automated Patch Clamp Studies

We studied 252 rare missense and in-frame indel *SCN5A* variants harbored by 458 of 3,335 individuals undergoing evaluation for BrS, described by Walsh et al.^12^ All experimental studies used the most common *SCN5A* transcript in the adult heart, which includes the adult isoform of exon 6 and a deletion of the alternatively spliced p.Gln1077 residue (ENST00000423572; MANE Select transcript). Our analyses use the nomenclature corresponding to this 2,015 residue isoform. However, we also present a second variant nomenclature corresponding to full 2,016 amino acid form that has often been used in the literature (ENST00000333535; Table S1). A single protein change (p.Asn1379Lys) was reported in two individuals, involving two different nucleotide changes (c.4137C>G and c.4137C>A). This variant was evaluated once in this study using the latter nucleotide change (c.4137C>A) (Table S1).

*SCN5A* variants were experimentally studied at Vanderbilt University Medical Center and/or Victor Chang Cardiac Research Institute as described in a recent publication.^19^ First, plasmids encoding inducible Na_V_1.5, recombination, and selection elements were independently generated by Quikchange mutagenesis (Agilent) or commercial oligonucleotide synthesis. Sequence-verified plasmids were then stably integrated into Human Embryonic Kidney (HEK293) cells. After selection for cells containing a single stable integration event, expression of Na_V_1.5 was induced. Cells were studied at each site using the SyncroPatch 384PE system (Nanion Technologies, Munich). All protocols for measuring Na_V_1.5 function (peak current density, pCD; voltages of steady state activation and inactivation, SSA and SSI; and recovery from inactivation, RFI) have been recently described.^19^ Analysis of experimental data was performed in R or MATLAB (see Data and Code Availability below).

The primary analysis used in this study involves the evaluation of pCD for variants measured after a step to -30 mV from a resting membrane potential of -90 mV, to mimic the physiologically-relevant cardiomyocyte membrane potential, as described by Ma et al.^19^ This ‘physiological’ (-90 mV) assay was updated and expanded in the present study to include data recorded for all control variants at both sites and updated mean and standard deviation (SD) ranges for the benign controls. We then calculated a *Z*-score for the ‘physiological’ pCD for each variant according to formula 1:

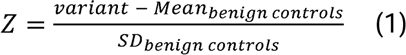

From this data *Z*-score ranges were defined as follows: *Z* >-2 defined as no LOF, -2 ≥ *Z* > -3: mild LOF, -3 ≥ *Z* > -4: moderate LOF; *Z* ≤ -4: severe LOF.

Independent site data were merged, and a weighted average was used for variants evaluated at both sites using formula 2:

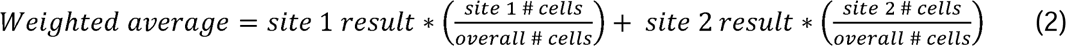

After application of the ClinGen Sequence Variant Interpretation Working Group recommendations,^21^ the updated assay had OddsPath scores of 23.96 and 0.043 for pathogenic and benign evidence, respectively (Figure S1; Table S2), equivalent to ACMG PS3_strong and BS3_strong evidence. However, we conservatively downgraded BS3_strong to BS3_moderate to account for potential functional mechanisms that were not captured by this assay.

We also performed a secondary analysis based on pCD and gating parameters. However, inclusion of gating parameters did not improve variant classifications or correlation with BrS penetrance so gating data were not used in the primary analysis.

*Cohort-level Case-Control Enrichment analysis.* Analyses of allele frequency used the precalculated gnomAD v4.1 filtering allele frequency (FAF) metric. FAF corresponds to the maximum allele frequency across major continental ancestries, with additional statistical adjustments.^22^ We defined rare variants as 0.001 > FAF > 0.00001 and ultra-rare variants as FAF<0.00001. The proportion of European ancestry BrS cases (n = 2,400) carrying rare variants was calculated for the entire *SCN5A* protein and for different topological regions. These regions were defined from a cryo-EM structure of Na_V_1.5 (PDB 6QLA),^23^ i.e., a) Pore loop regions I-IV: residues 273-389, 862-912, 1356-1444, 1679-1743; b) Transmembrane regions (non-pore loops) I-IV: residues 132-410, 718-938, 1206-1469, 1528-1771 (excluding pore loop region residues); c) N-terminus: residues 1-131; d) Interdomain linker (IDL) regions: residues 411-717, 939-1205, 1470-1527; and e) C-terminus: residues 1772-2015. For each region, the proportion of cases carrying ultra-rare or rare variants was sub-classified according to the functional ACMG (PS3/BS3) evidence applied or the overall ACMG classification. Control frequencies were calculated using the non-Finnish European (NFE) samples of the gnomAD v3 genomes dataset as it has a better breadth of coverage with WGS compared to WES for calculating overall control frequencies.

### ACMG Criteria Implementation and Evaluation

Variant functional data (PS3/BS3) were assigned using our *Z*-score approach.^19, 24^ Variants were binned by *Z*-score according to degree of LOF, ranging from PS3 to BS3_moderate: PS3_strong: *Z* ≤ -4; PS3_mod -4 < *Z* ≤ -3; PS3_supp -3 < *Z* ≤ -2; BS3_supp -2 < *Z* ≤ -1; BS3_mod *Z* > -1. For variant AF, we used population data from gnomAD v4.1,^22^ implementing PM2_supporting (FAF < 0.00003) and BS1 (max population AF > 0.0003).^19^ Case enrichment (PS4) for single variants was implemented as described by Walsh et al.^12^ In-frame indel variants received PM4 evidence due to a change in protein length. The hot-spot criterion (PM1) was assessed based on location within the protein and gnomAD AF stratified by genetic ancestry (European vs East Asian; see differential *SCN5A*-BrS burden in Walsh et al)^12^ to provide up to moderate level evidence. Two *in silico* predictors were used to apply computational variant effect criteria (PP3/BP4): REVEL^15^ and AlphaMissense.^14^ We present sensitivity analyses of each variant effect predictor by limiting implementation to (1) only supporting or (2) allowing strong by ‘capping’ combined PP3/PM1 criteria to strong based on a previous calibration.^25^ Thresholds for each criterion implementation are outlined in Table S3. Each criterion strength was assigned a point value according to the ACMG Bayesian points-based system.^26^ Criteria were combined by adding or subtracting points for eventual classification. Under the Bayesian point scale system,^26^ final point scores ≥ 10 or ≥ 6 were required for pathogenic classifications P or LP, respectively, and final point scores ≤ -1 or ≤ -7 were required for benign classifications LB or B, respectively. However, to classify variant as LB, we used a more conservative cutoff of -5 points, which corresponds to 1 strong benign and 1 supporting benign criteria, matching the 2015 ACMG/AMP guidelines (Table S4)^10^.

*Splicing.* We obtained SpliceAI scores^27^ from the SpliceAI website (https://spliceailookup.broadinstitute.org/), using default parameters (hg38, Gencode basic, max distance 500 bp, no masked scores). Aggregate SpliceAI scores were calculated using the formula:

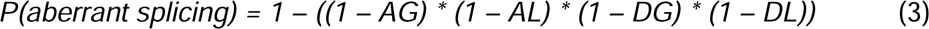

where AG, AL, DG, and DL indicate scores for Acceptor Gain, Acceptor Loss, Donor Gain, and Donor Loss, respectively. Because the primary focus of this paper is to determine the predictive ability of the APC assay, we present the SpliceAI scores in Table S5 but did not integrate them towards variant classification.

*SCN5A Variant Brugada Syndrome Penetrance Estimates.* Genotype-first *SCN5A*-BrS penetrance estimates were calculated using a Bayesian binomial framework, adapting a recently described phenotype-agonistic method by McGurk et al.^28^ This strategy specifically addressed penetrance among variants discovered as ‘secondary findings’, rather than penetrance within family studies/ascertained sequencing. Penetrance estimates and confidence intervals were calculated using the *penetrance* R function obtained from https://github.com/ImperialCardioGenetics/variantfx/tree/main/PenetrancePaper. Specifically, we predicted *SCN5A*-BrS_penetrance as:

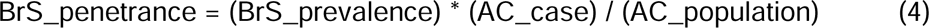

Where allele counts (AC) are obtained from the BrS cohort above (AC_case), a population estimate from gnomAD (AC_population), and an estimate of previous BrS prevalence (1:2000 based on a meta-analysis of 26 studies spanning 388,237 individuals)^7, 29^. The above model explicitly incorporates epidemiological data and relevant differences in AC ascertainment to account for penetrance within different study applications. Notably, the case definition for this referral cohort was focused on EKG changes with sodium channel blocker provocation, which may not fully capture a pure BrS phenotype. Next, comparisons of variant penetrance for groups of *SCN5A* variants were assessed across ranges of several predictor scores/classifications: AlphaMissense score (using calibrated score ranges)^16^, REVEL score (using calibrated score ranges).^16^ hot-spot criteria (PM1_mod, PM1_supp, or none, as described above), pCD *Z*-score (in bins of width 1), pCD ACMG criterion (as described above), and ACMG classification. For each bin, we determined total BrS cases,^12^ gnomAD allele count, average AF for cases, and average AF of gnomAD variants.^22^

### Statistical Analysis

For variant APC analyses, biological and technical replicates were performed per variant and experiment, respectively. Following our previous power calculation, a minimum of 36 cells per variant were measured.^19^ *SCN5A*-BrS odds ratios were determined for aggregate groups of variants based on functional data and variant classifications. Across each group, the odds ratios were calculated using formula 4 below. Control counts were taken from gnomAD v4.1 and total BrS cases from the BrS cohort.^12^

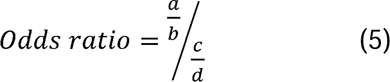

where, a = count of BrS cases across the group of variants, , = count of BrS cases without the variants, , = count of gnomAD with variants, d = count of gnomAD without variants.

## Results

### Comprehensive SCN5A APC Data for a BrS Cohort

We employed our calibrated multi-site APC assay to quantify the functional effect of 252 missense and in-frame indel *SCN5A* variants in HEK293 cells (Figure 1A and Table S6).^19^ We first measured Na_V_1.5 sodium pCD, the primary molecular correlate of *SCN5A*-BrS, from a holding potential of -90 mV. Example traces are shown in Figure 1B, and pCD measurements for all variants are presented in Table S6. Reduced pCD was observed in 146 variants (58%). This includes 100 variants (40%) with severe LOF, 25 (10%) with moderate LOF and 21 (8%) with mild LOF, corresponding to PS3_strong, PS3_moderate and PS3_supporting, respectively. Conversely, 106 variants (42%) had peak currents within the normal range that conferred BS3_moderate or BS3_supporting evidence (Figure 1C). We also recorded gating properties (voltage of half activation, voltage of half inactivation, and recovery from inactivation) if there was sufficient current (Table S6). Due to its strong ability to distinguish control benign from pathogenic variants (Figure 1C),^19^ we used pCD as the primary functional score for subsequent analyses. Six missense variants in the cohort had aggregate SpliceAI scores above 0.5 and 20 missense variants had scores between 0.2 and 0.5 (Table S5). Of these 26 variants, 10 had current densities in the normal range (*Z* > - 2) and thus are candidates to disrupt Na_V_1.5 function primarily through a splicing mechanism,^30^ which is not captured by this APC assay.

**Figure 1.**
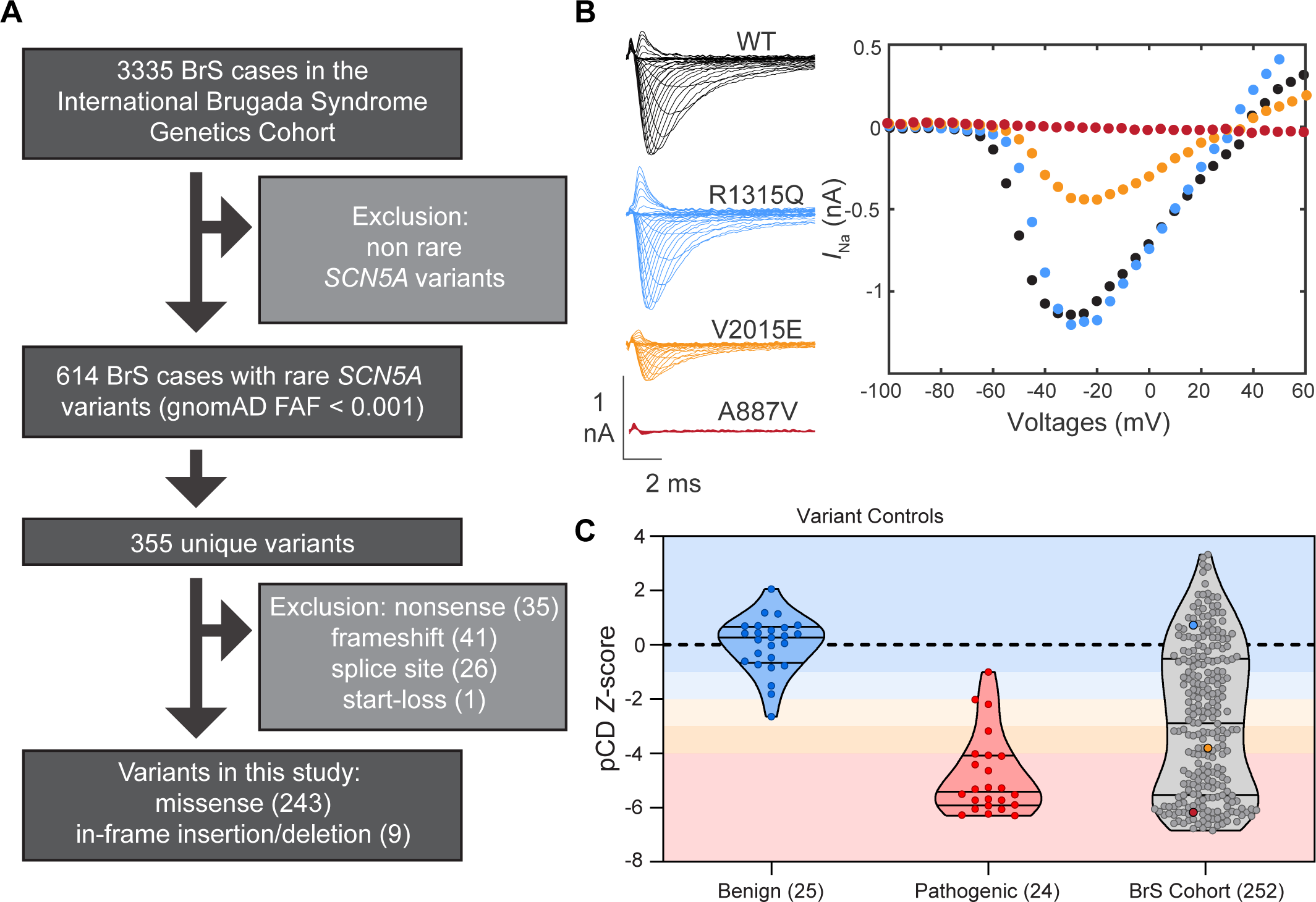
APC of *SCN5A* variants detected in a cohort of patients with Brugada Syndrome (BrS). **A**. Overview of BrS cohort described in Walsh et al., 2021. The cohort includes 252 unique missense and in-frame indel variants were analyzed on the protein level. These variants account for 458 of the 614 BrS cases involving rare *SCN5A* variants. **B**. Example *I*_Na_ traces holding at -90 mV and stimulating at voltages ranging from -100 mV to +60 mV in 5 mV intervals. Traces and current-voltage curves are shown for WT and R1315Q, V2015E and A887V, which had *Z*-scores of 0.73, -3.82 and -6.17, corresponding to BS3_moderate, PS3_moderate and PS3_strong, respectively. **C**. *Z*-score of peak current density (pCD) for benign and pathogenic variant controls and the BrS cohort of 252 variants. Colored circles in the BrS cohort correspond to example traces R1315Q (blue), V2015E (yellow) and A887V (red).

### Functional Assessment by Protein Region

We next stratified variant functional scores across Na_V_1.5 regions: pore loop, transmembrane (non-pore loop), N-terminus, C-terminus, and interdomain linker (IDL) regions (defined in the Methods). Most missense and in-frame indel variants in this cohort were in transmembrane regions (Figure 2A-C; N = 184/252, 73.0%). The transmembrane pore loop regions showed the highest fraction of LOF variants: 80/94 variants had LOF scores (*Z* ≤ -2). In the transmembrane domains outside the pore loop region, 49/90 variants were LOF. Variants within the IDL and C-terminus regions had the highest fractions of functionally normal variants at 31/38 and 13/18 with *Z* > -2, respectively (Figure 2D). Despite the enrichment of LOF variants in the transmembrane regions, there were nonetheless 55 rare and ultra-rare variants with normal function in these hot-spot domains (Figure 2D). An example of normal-function variant in a hot-spot region is p.Ala364Val in the DI pore loop (BS3_moderate, *Z* = -0.07; present in a single BrS case in the BrS cohort and a single participant in gnomAD v4.1) which also is predicted to have normal splicing by SpliceAI (Table S5).

**Figure 2:**
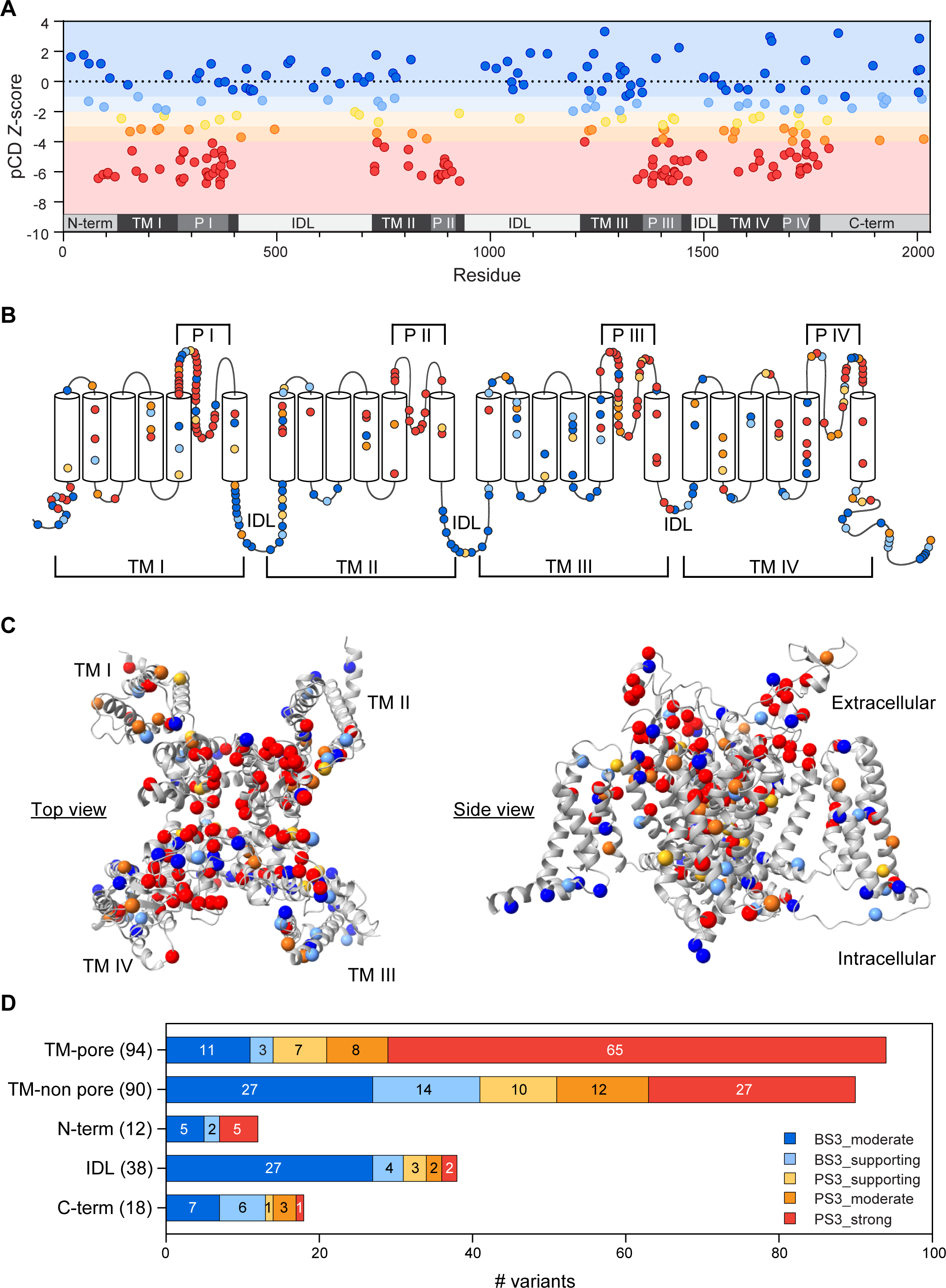
Functional scores of *SCN5A*-BrS variants by protein domain. **A**. *Z*-scores of variants in the cohort plotted against position in the protein. **B.** Two-dimensional topological view of Na_V_1.5. **C**. Three-dimensional structure of hNa_V_1.5 (PDB: 6LQA^23^). Many N-term, IDL, and C-term variants are not present in the structure and are omitted from this plot. **D**. Counts of functional criteria by location in protein. **A-D.** Variants are color-coded by ACMG classification level as defined in the legend in the lower right. Most variants found in the BrS cases are in the transmembrane and pore loop regions, and these regions are also enriched for loss-of-function variants. N-term indicates N-terminus; TM, transmembrane; P, pore; IDL, inter-domain linker; C-term, C-terminus; pCD, peak current density.

### Variant Classification without and with APC Data

We evaluated the impact of the 252-variant APC dataset on variant classifications by combining the functional data with other criteria, including hot-spot, AF, case counts, and computational predictors (Table S7). Two computational tools, REVEL^15^ and AlphaMissense^14^ were used. We assessed variant classifications using REVEL and AlphaMissense with ClinGen-recommended calibration cutoffs and classification guidelines.^16, 25^ Variants were classified with or without functional data. Using calibrated REVEL, 220 variants were classified as VUS and 32 as LP at baseline. Including APC data led to 2 LB, 113 VUS, 118 LP, and 19 P classifications (Figure 3A-B). For AlphaMissense, baseline classifications were 3 LB, 225 VUS and 24 LP. Adding APC data revised these to 2 B, 7 LB, 117 VUS, 111 LP, and 15 P (Figure 3C-D). These findings demonstrate that incorporating functional data enables significant variant reclassification, with many VUS shifting to LP and LP to P, across various classification rules. We also assessed variant classifications using *in silico* evidence at a supporting level, as initially recommended by 2015 ACMG criteria, which enabled the reclassification of up to 99 VUS (Figure S2, Table S8).

**Figure 3:**
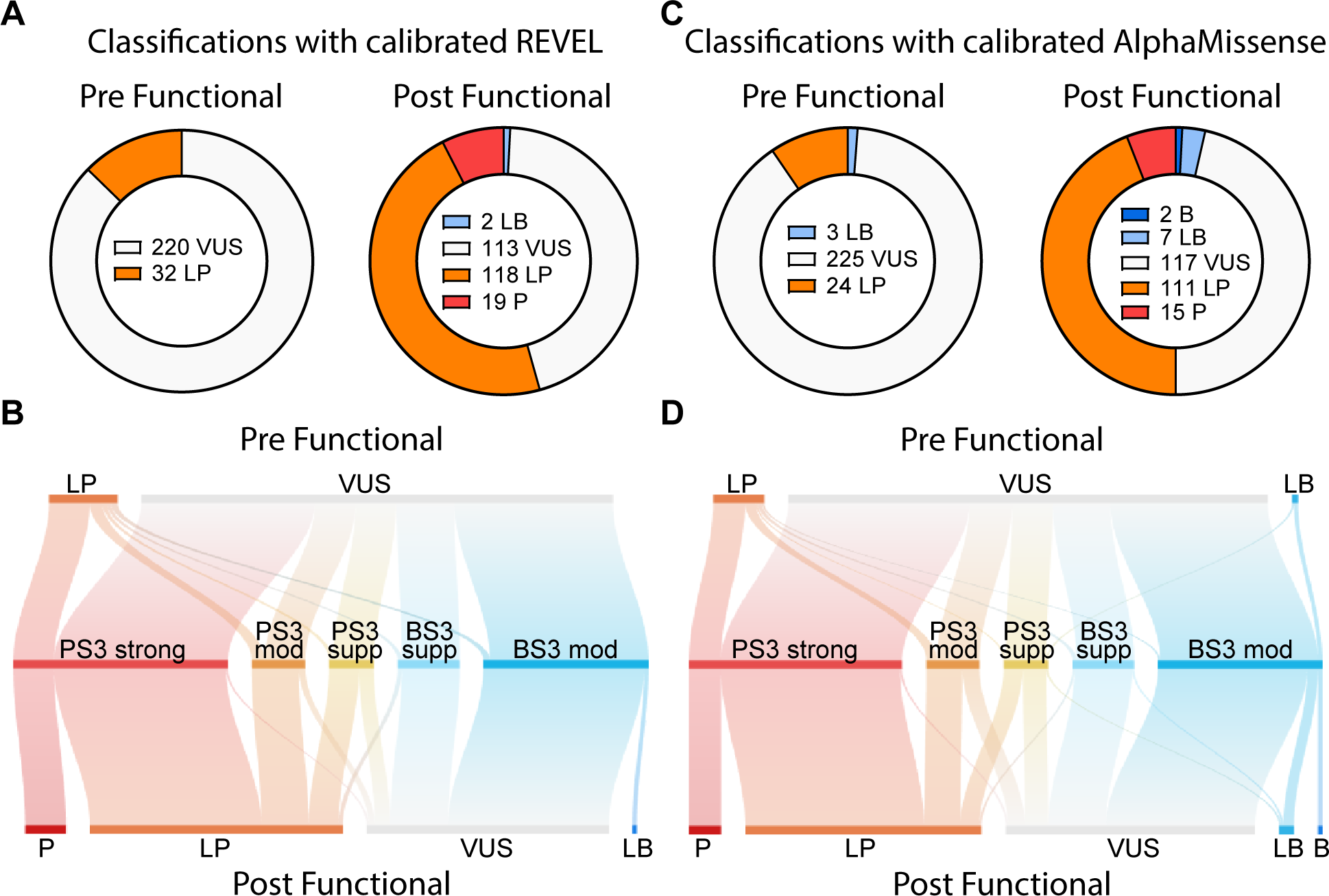
Impact of functional data (PS3/BS3) on variant classifications. Variant classifications for 252 *SCN5A* variants from the BrS cohort were pre- and post- applying functional APC evidence (PS3_strong to BS3_moderate). Variants were classified using population frequency (BS1), variant rarity (PM2_supporting), hot-spot evidence (PM1 at a maximum of moderate), in-frame indel (PM4) and case enrichment (PS4). **A** through **D,** Calibrated *in silico* predictions (PP3/BP4) from REVEL (**A**, **B**) or AlphaMissense (**C**, **D**) were applied, at a maximum level of 4 points when combined with PM1. Sankey plots show flow between functional evidence and variant classifications for calibrated REVEL (**B**) and calibrated AlphaMissense (**D**). A full list of criteria is provided in Table S3. Figure S2 shows a similar analysis, but with the application of REVEL and AlphaMissense at a maximum of a supporting level.

### Enrichment of Affected BrS heterozygotes by Protein Region

Case-control studies have consistently noted a strong enrichment of ultra-rare (gnomAD FAF<0.00001) missense variants in *SCN5A* in BrS cohorts compared to control or population datasets.^12, 31^ The proportion of BrS cases and gnomAD controls with ultra-rare missense variants, for the entire protein and the major protein regions are shown in Figure 4A. The proportion of BrS cases with LOF variants (PS3_supporting to strong rule applied) falls comfortably within the case excess for the overall analysis and the five regions separately, indicating that the assay is not generally overestimating reduced sodium channel function or variant pathogenicity. It is also important to note that the vast majority of LOF variants fall into the ultra-rare category. A more marginal excess of rare (0.001 > gnomAD v4.1 FAF > 0.00001) *SCN5A* missense variants is also observed in BrS cases, which is entirely restricted to the transmembrane region (for these European-ancestry cases) (Figure 4B). Overall, 9.25% of patients in the BrS cohort had an ultra-rare LOF (PS3 supporting to strong) variant and 0.46% had a rare LOF variant. For BrS patients with an ultra-rare *SCN5A* variant, 77.6% of the time the variant had LOF. However, for BrS patients with a rare *SCN5A* variant, only 13.6% of the time the variant had LOF.

**Figure 4:**
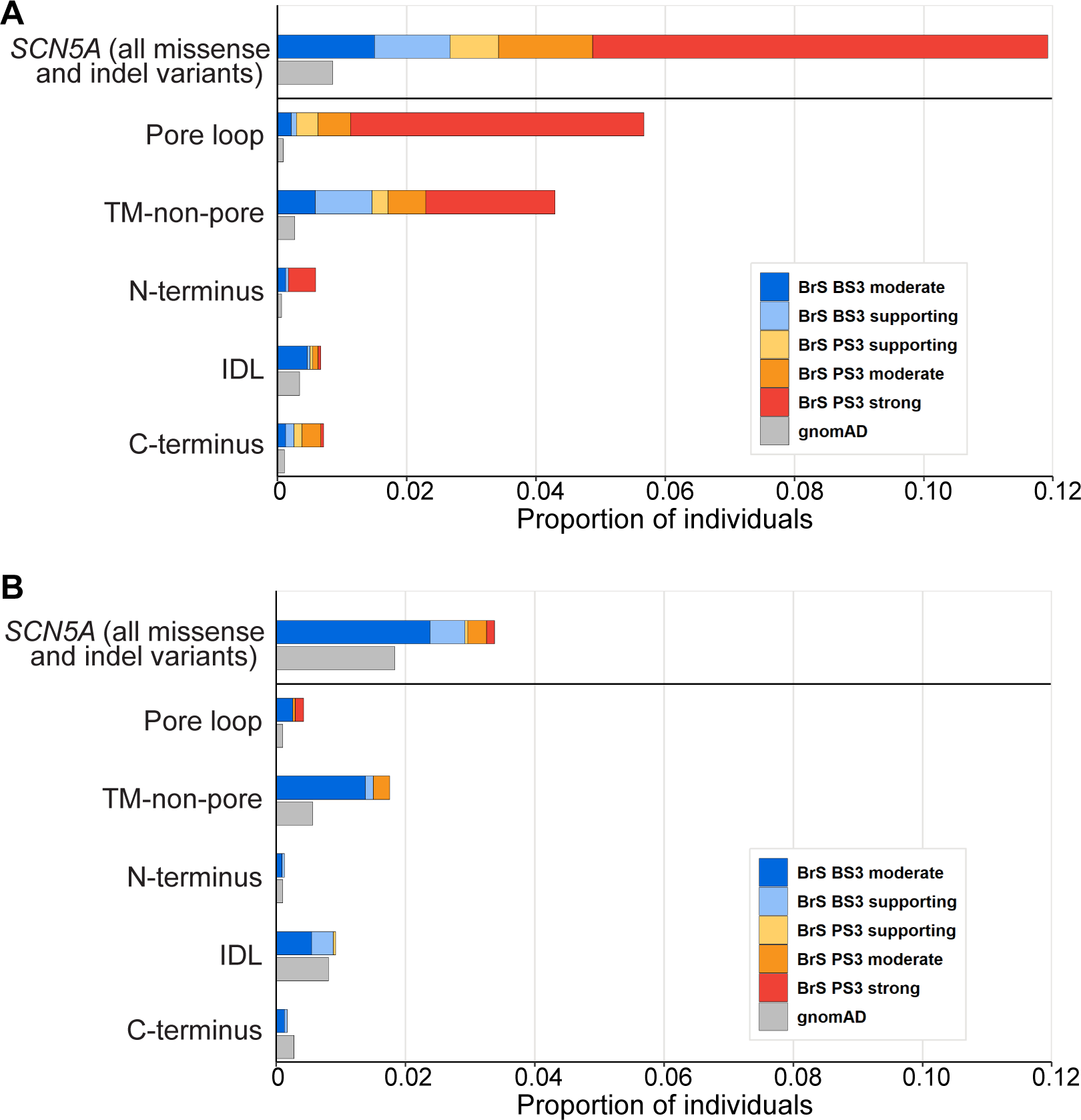
Proportion of individuals that have ultra-rare or rare *SCN5A* variants by protein region. **A**. Ultra-rare variants (gnomAD v4.1 FAF <0.00001) are located mostly in the transmembrane (TM) domains and are functionally abnormal except for the few in the Inter- domain Linker (IDL) region. **B**. Rare variants with a gnomAD v4.1 FAF between 0.00001 and 0.001 are mostly functionally normal in all domains.

### Cohort Variant Brugada Syndrome Penetrance

Like many dominant Mendelian disease genes, *SCN5A* variants associated with BrS have incomplete penetrance.^8^ We therefore investigated whether our high-throughput functional data could improve stratification of variant-specific penetrance. We first assessed aggregate BrS penetrance and odds ratios for BrS by variant classification. The complete set of penetrance and odds ratio values are presented in Figure 5 and Table S9. For the final ACMG classifications (using calibrated AlphaMissense), P variants had a penetrance of 0.18 (odds ratio of 371), LP variants had a penetrance of 0.09 (OR 185), and VUS, LB, and B variants each had a penetrance below 0.01 (Figure 5A, see Figure S3 for final ACMG classification using calibrated REVEL and *in silico* at supporting levels). We next examined the impact of individual ACMG criteria for stratifying BrS risk. Calibrated AlphaMissense and REVEL (when used alone) had a moderate ability to stratify BrS risk, with a maximum penetrance of 0.08 (OR 169) for AlphaMissense PP3_strong and 0.06 (OR 129) for REVEL PP3_strong (Figure 5B-C). Similarly, hot-spot evidence (when used alone) had moderate predictive ability, with a maximum penetrance of 0.08 (OR 165) for BrS for PM1_moderate (Figure 5D). Functional data had the strongest predictive ability of any single criterion. When binned by ACMG evidence level, functional data had a maximum penetrance of 0.15 (OR 318) for PS3_strong (Z ≤ -4; Figure 5E). When more granularly binned by Z-score, functional data had an even higher predictive ability, with a maximum penetrance of 0.25 (OR 501) for Z ≤ -6 (Figure 5F).

**Figure 5:**
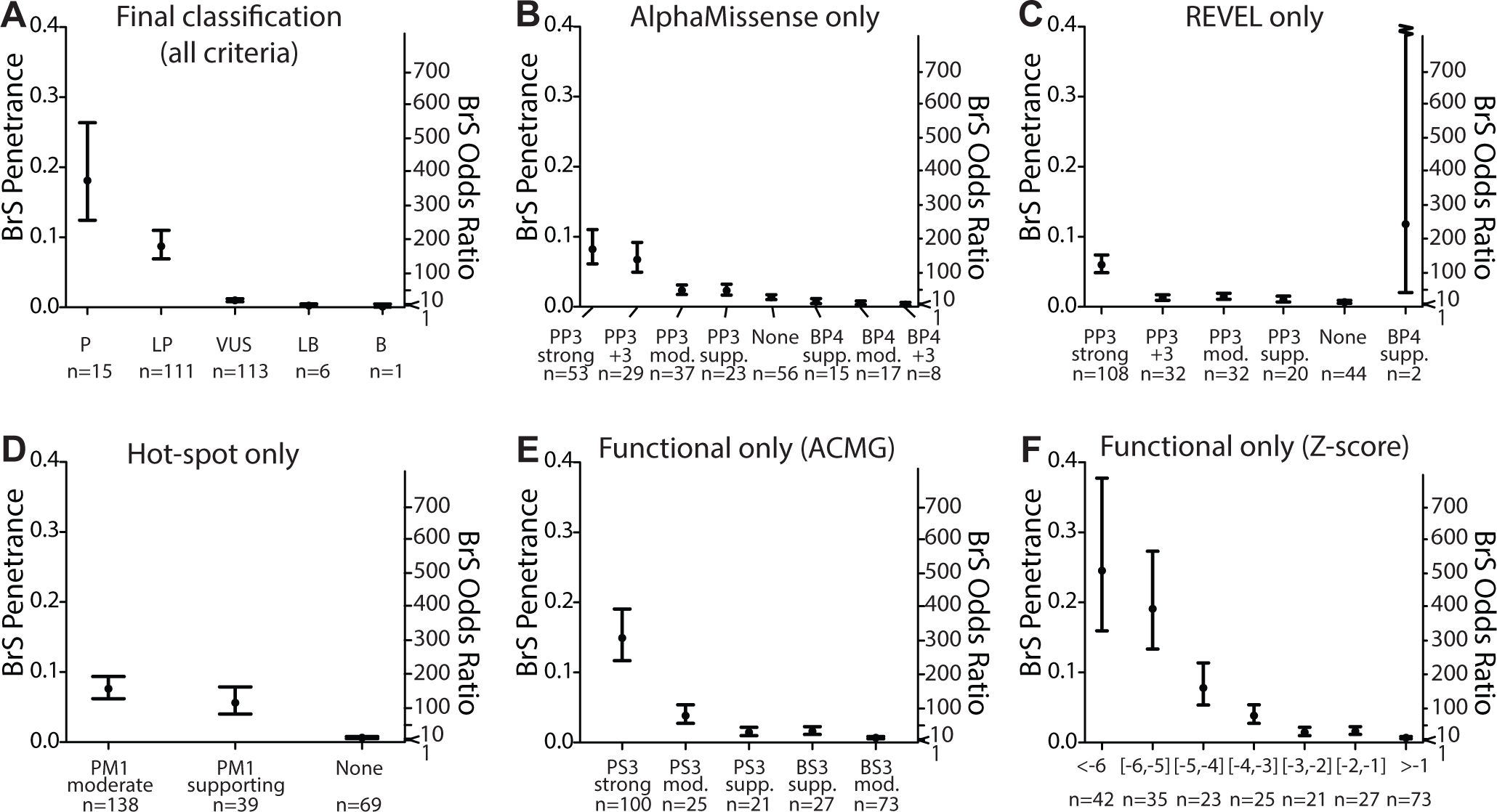
BrS Penetrance and Odds Ratio based on classifications or variant features. Variants were stratified by final classification (integrating all criteria including calibrated AlphaMissense) (**A**), calibrated AlphaMissense (**B**), calibrated REVEL (**C**), hot-spot evidence (**D**), functional evidence (using ACMG evidence categories) (**E**) or functional evidence derived from pCD *Z*-score (**F**). Figure S3 shows the penetrance stratified by final classification using calibrated REVEL, and supporting AlphaMissense and REVEL.

## Discussion

*SCN5A* LOF variants are a major monogenic cause of BrS.^32, 33^ However, many *SCN5A* missense variants are classified as VUS due to limited available evidence. In this study, we assessed *SCN5A* variants identified from a large cohort of BrS patients^12^ using a high-throughput APC assay that we have previously validated.^19^ Of 252 tested variants, 146 had LOF, including 100 with strong LOF. Incorporating this functional evidence with other classification criteria resulted in reclassification of ∼48% of VUS. Furthermore, the degree of functional perturbation significantly corelated with BrS OR in a clinical cohort, and penetrance for a secondary-finding population, respectively.

Our dataset is the most extensive collection of *SCN5A* variant functional data to date. Of the 252 rare and ultra-rare missense and in-frame indel variants studied, we identified 146 variants with loss of Na_V_1.5 function observed through reduced pCD, the primary molecular correlate of BrS.^18, 34^ LOF variants were enriched in transmembrane domains, especially the pore loop region; individuals harboring transmembrane domain variants have been shown to experience worse outcomes.^33^ LOF variants were also enriched for ultra-rare variants (gnomAD FAF<0.00001), compared to rare variants (FAF between 0.00001 and 0.001). This data, together with previous mathematical analyses^35^ and case-control data,^12^ support a model where most of the *SCN5A* large-effect BrS burden is concentrated among the ultra-rare variants, as opposed to the rare variants. Interestingly, 106 rare variants found in BrS patients exhibited normal function, with 77 categorized as BS3_moderate and 29 as BS3_supporting. Most of these functionally normal variants are located outside the “hot-spot” transmembrane domains and most have allele frequencies in the rare range (0.001 > gnomAD FAF > 0.00001) compared to “ultra-rare” variants (gnomAD FAF<0.00001). The residual excess of variants with benign functional evidence in the BrS cohort compared to controls suggests that a subset of these variants may also be contributing to BrS pathophysiology. These variants could be affecting sodium channel function through mechanisms not evaluated by our HEK cell APC assay (*e.g.* by disrupting splicing). These variants could also have modest effect sizes insufficient to achieve a pathogenic functional status, particularly those variants with a BS3_supporting classification. However, as a group, these ‘normal-function’ variants show much lower BrS penetrance than LOF variants, suggesting that most are “bystander” variants that do not strongly contribute to arrhythmias.^32, 33^

We used the APC data to help reclassify the 252 *SCN5A* variants using the ACMG/AMP framework. Although the framework has greatly improved the consistency of variant classifications, there are still debates and inconsistent application of varying evidence strengths and rules for variant classification.^36, 37^ For example, computational evidence (PP3), originally capped at supporting strength,^10^ has recently been calibrated by the ClinGen Sequence Variant Interpretation working group to provide up to strong evidence for classification.^16, 25^ We therefore conducted contingency analyses using two *in silico* predictions (REVEL^15^ and AlphaMissense^14^) applied at two different evidence strengths (maximum of supporting^10^ or using recent ClinGen-calibrated cutoffs^16^). Our analysis showed that combining functional evidence with these *in silico* tools enables reclassification of 99 or 98 abnormal variants from VUS to likely pathogenic when *in silico* prediction was applied at supporting for REVEL or AlphaMissense, respectively. This number further increased to 108 or 104 when calibrated *in silico* predictions were used. Our functional data also prevented 14 or 7 functionally normal VUS from being reclassified to LP due to strong-level pathogenic predictions from REVEL or AlphaMissense, respectively. Lastly, our normal functional evidence helped reclassify 2 and 6 rare VUS to LB when calibrated REVEL and AlphaMissense were used, respectively. Notably, AlphaMissense performed better than REVEL in identifying functionally normal variants as “benign” (Figure 4, Figure S4). Overall, functional data enabled reclassification of 48.6% of VUS when using calibrated REVEL and 48.0% of VUS when using calibrated AlphaMissense. We note that since we studied variants detected in patients with BrS, there was an ascertainment bias that increased the prevalence of deleterious variants compared to the study of “randomly” occurring variants or variants detected through population screening.^38^ Notably, of 146 LOF variants, 46 have applicable PS3_moderate or supporting evidence but 10/38 and 19/39 remained as VUS when using our classification strategy with calibrated REVEL or AlphaMissense, respectively. Our abnormal functional evidence will aid in the reclassification of these VUS in the future if more patients are identified with these VUS. There is also an asymmetry where our framework is more likely to reclassify variants from VUS to LP than from VUS to LB. This largely results from our conservative capping of normal functional evidence at BS3_moderate and the use of a more stringent threshold of -5 points to reach LB. As variant classification schemes continue to evolve, our *SCN5A* functional data will be integrated to further improve classification of *SCN5A* variants.

The incomplete penetrance of LOF variants in “Mendelian” disease genes has been increasingly recognized, especially with the availability of relatively unselected population and biobank cohorts.^22, 28, 39^ For BrS, only about 20% of BrS patients have rare *SCN5A* variants, and these variants furthermore typically exhibit incomplete penetrance.^40^ There are numerous examples of *SCN5A* variants causing variable phenotypes, including no phenotype, within a single family.^41–44^ In addition to penetrance within family studies and ascertained clinical workup, genotype-first precision medicine will increasingly rely on adjudication of variants observed in unascertained populations, such as those involved as secondary-findings.^11^ Previous work on *SCN5A* variant penetrance has focused on penetrance-estimates incorporating proactive integration of variant features using an expectation-maximization framework integrating several study designs^40, 45^. Here, we used an approach established by McGurk et al. to use binned sets of variant features (including functional data) or variant classifications to estimate penetrance in a unascertained population.^28^ Although many variant features correlated with BrS penetrance, severity of LOF in the APC assay was the feature that best correlated with risk for BrS. These data suggest that when APC data are available, they could be used to inform the value of return of secondary-findings, an increasingly important clinical challenge.^11^ This analysis specifically focuses on *SCN5A*-BrS penetrance, which only partially captures the genetic architecture of BrS. We note that manifestation of BrS is likely modulated by many features not modelled in this study, including demographics, environmental influences, polygenic pathways,^9, 46, 47^ and/or yet-to-be-identified monogenic influences.^48^

## Limitations

Although *SCN5A* is a pleiotropic gene and *SCN5A* variants can cause a variety of arrhythmia and heart failure phenotypes (including Long QT Type 3, Dilated Cardiomyopathy, Multiple Ectopic Premature Purkinje Contractions), our functional assay is currently calibrated specifically for the *SCN5A*-BrS relationship. The frequency of *SCN5A* LOF variants is reported to differ among distinct genetic ancestry groups^49^, and this analysis includes few cases of non-European and non-East Asian ancestry. While patient demographics and common genetic background influence BrS risk, we could not account for these variables within the available data. Our assay used a cDNA form of *SCN5A* that cannot model variant effects on splicing. Variants found in BrS patients with moderate or high SpliceAI scores can be further studied with complementary splicing assays, as we have recently demonstrated.^30, 50^ Penetrance estimates are limited by varied allele frequencies in referral populations and phenotype definitions (ECG findings as criteria in the BrS cohort).

## Conclusions

In this study, we deployed a calibrated functional assay on 252 rare missense and in-frame indel variants detected in a large BrS cohort. Integration of this data with AF, case-enrichment, hot-spot, and computational evidence enabled the reclassification of approximately 48% of VUS according to ACMG criteria. We also demonstrated a strong correlation between the extent of LOF and variant penetrance. Our dataset and approach should further aid in the clinical diagnosis and management of probands and their families.

## Data Availability and Code Availability

All experimental and *in silico* data generated and used in this analysis are included in this study. Code to analyze these data are available at the Glazer Lab GitHub site: https://github.com/GlazerLab/SCN5A_SP_BrS_cohort and at the VCCRI GitHub site: https://github.com/VCCRI/SCN5A_SyncroPatch. Variant functional data and classifications will be submitted to ClinVar upon acceptance of the manuscript. All included patient data were derived from previously published, deidentified data.

## Acknowledgements

We thank Kenneth Matreyek and Doug Fowler for the HEK293 landing pad cells, and Margaret L. Harvey and Maria Calandranis for *SCN5A* cloning assistance and cell preparation. We also acknowledge support from the Victor Chang Cardiac Research Institute Innovation Centre, funded by the NSW Government.

---

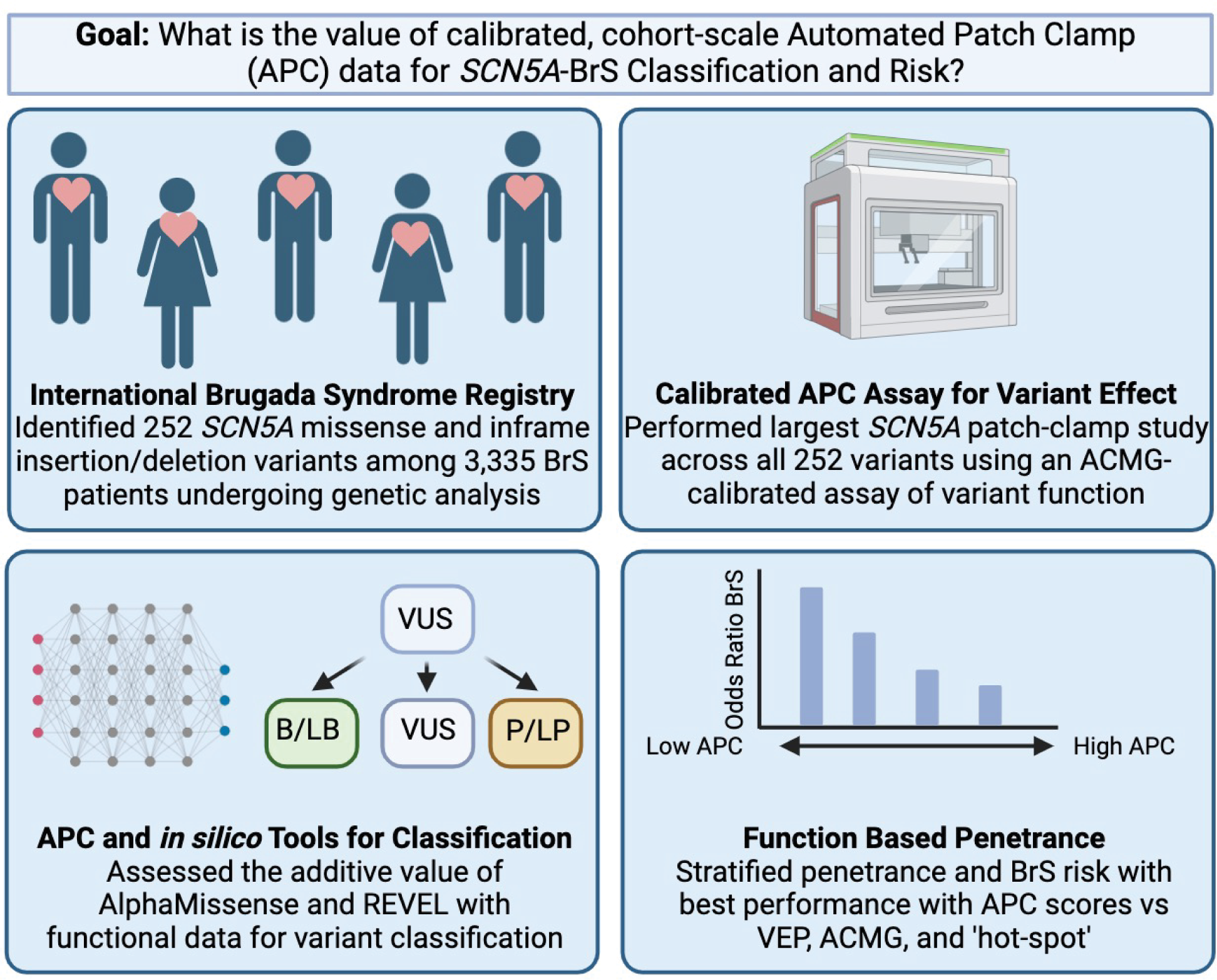

## References

1. Cerrone M, Costa S and Delmar M. The Genetics of Brugada Syndrome. Annu Rev Genomics Hum Genet. 2022.

2. Milman A, Andorin A, Gourraud JB, Postema PG, Sacher F, Mabo P, Kim SH, Juang JJM, Maeda S, Takahashi Y, Kamakura T, Aiba T, Conte G, Sarquella-Brugada G, Leshem E, Rahkovich M, Hochstadt A, Mizusawa Y, Arbelo E, Huang Z, Denjoy I, Giustetto C, Wijeyeratne YD, Napolitano C, Michowitz Y, Brugada R, Casado-Arroyo R, Champagne J, Calo L, Tfelt-Hansen J, Priori SG, Takagi M, Veltmann C, Delise P, Corrado D, Behr ER, Gaita F, Yan GX, Brugada J, Leenhardt A, Wilde AAM, Brugada P, Kusano KF, Hirao K, Nam GB, Probst V and Belhassen B. Profile of patients with Brugada syndrome presenting with their first documented arrhythmic event: Data from the Survey on Arrhythmic Events in BRUgada Syndrome (SABRUS). Heart Rhythm. 2018;15:716–724.

3. Brugada P and Brugada J. Right bundle branch block, persistent ST segment elevation and sudden cardiac death: a distinct clinical and electrocardiographic syndrome. A multicenter report. J Am Coll Cardiol. 1992;20:1391–6.

4. Dereci A, Yap SC and Schinkel AFL. Meta-Analysis of Clinical Outcome After Implantable Cardioverter-Defibrillator Implantation in Patients With Brugada Syndrome. JACC Clin Electrophysiol. 2019;5:141–148.

5. Hosseini SM, Kim R, Udupa S, Costain G, Jobling R, Liston E, Jamal SM, Szybowska M, Morel CF, Bowdin S, Garcia J, Care M, Sturm AC, Novelli V, Ackerman MJ, Ware JS, Hershberger RE, Wilde AAM and Gollob MH. Reappraisal of Reported Genes for Sudden Arrhythmic Death: Evidence-Based Evaluation of Gene Validity for Brugada Syndrome. Circulation. 2018;138:1195–1205.

6. Wilde AAM and Amin AS. Clinical Spectrum of SCN5A Mutations: Long QT Syndrome, Brugada Syndrome, and Cardiomyopathy. JACC Clin Electrophysiol. 2018;4:569–579.

7. Mizusawa Y and Wilde AA. Brugada syndrome. Circ Arrhythm Electrophysiol. 2012;5:606–16.

8. Wijeyeratne YD, Tanck MW, Mizusawa Y, Batchvarov V, Barc J, Crotti L, Bos JM, Tester DJ, Muir A, Veltmann C, Ohno S, Page SP, Galvin J, Tadros R, Muggenthaler M, Raju H, Denjoy I, Schott JJ, Gourraud JB, Skoric-Milosavljevic D, Nannenberg EA, Redon R, Papadakis M, Kyndt F, Dagradi F, Castelletti S, Torchio M, Meitinger T, Lichtner P, Ishikawa T, Wilde AAM, Takahashi K, Sharma S, Roden DM, Borggrefe MM, McKeown PP, Shimizu W, Horie M, Makita N, Aiba T, Ackerman MJ, Schwartz PJ, Probst V, Bezzina CR and Behr ER. SCN5A Mutation Type and a Genetic Risk Score Associate Variably With Brugada Syndrome Phenotype in SCN5A Families. Circ Genom Precis Med. 2020;13:e002911.

9. Barc J, Tadros R, Glinge C, Chiang DY, Jouni M, Simonet F, Jurgens SJ, Baudic M, Nicastro M, Potet F, Offerhaus JA, Walsh R, Choi SH, Verkerk AO, Mizusawa Y, Anys S, Minois D, Arnaud M, Duchateau J, Wijeyeratne YD, Muir A, Papadakis M, Castelletti S, Torchio M, Ortuño CG, Lacunza J, Giachino DF, Cerrato N, Martins RP, Campuzano O, Van Dooren S, Thollet A, Kyndt F, Mazzanti A, Clémenty N, Bisson A, Corveleyn A, Stallmeyer B, Dittmann S, Saenen J, Noël A, Honarbakhsh S, Rudic B, Marzak H, Rowe MK, Federspiel C, Le Page S, Placide L, Milhem A, Barajas-Martinez H, Beckmann BM, Krapels IP, Steinfurt J, Winkel BG, Jabbari R, Shoemaker MB, Boukens BJ, Škorić-Milosavljević D, Bikker H, Manevy FC, Lichtner P, Ribasés M, Meitinger T, Müller-Nurasyid M, Veldink JH, van den Berg LH, Van Damme P, Cusi D, Lanzani C, Rigade S, Charpentier E, Baron E, Bonnaud S, Lecointe S, Donnart A, Le Marec H, Chatel S, Karakachoff M, Bézieau S, London B, Tfelt-Hansen J, Roden D, Odening KE, Cerrone M, Chinitz LA, Volders PG, van de Berg MP, Laurent G, Faivre L, Antzelevitch C, Kääb S, Arnaout AA, Dupuis JM, Pasquie JL, Billon O, Roberts JD, Jesel L, Borggrefe M, Lambiase PD, Mansourati J, Loeys B, Leenhardt A, Guicheney P, Maury P, Schulze-Bahr E, Robyns T, Breckpot J, Babuty D, Priori SG, Napolitano C, de Asmundis C, Brugada P, Brugada R, Arbelo E, Brugada J, Mabo P, Behar N, Giustetto C, Molina MS, Gimeno JR, Hasdemir C, Schwartz PJ, Crotti L, McKeown PP, Sharma S, Behr ER, Haissaguerre M, Sacher F, Rooryck C, Tan HL, Remme CA, Postema PG, Delmar M, Ellinor PT, Lubitz SA, Gourraud JB, Tanck MW, George AL, Jr., MacRae CA, Burridge PW, Dina C, Probst V, Wilde AA, Schott JJ, Redon R and Bezzina CR. Genome-wide association analyses identify new Brugada syndrome risk loci and highlight a new mechanism of sodium channel regulation in disease susceptibility. Nat Genet. 2022;54:232–239.

10. Richards S, Aziz N, Bale S, Bick D, Das S, Gastier-Foster J, Grody WW, Hegde M, Lyon E, Spector E, Voelkerding K and Rehm HL. Standards and guidelines for the interpretation of sequence variants: a joint consensus recommendation of the American College of Medical Genetics and Genomics and the Association for Molecular Pathology. Genet Med. 2015;17:405–24.

11. Miller DT, Lee K, Abul-Husn NS, Amendola LM, Brothers K, Chung WK, Gollob MH, Gordon AS, Harrison SM, Hershberger RE, Klein TE, Richards CS, Stewart DR and Martin CL. ACMG SF v3.2 list for reporting of secondary findings in clinical exome and genome sequencing: A policy statement of the American College of Medical Genetics and Genomics (ACMG). Genet Med. 2023;25:100866.

12. Walsh R, Lahrouchi N, Tadros R, Kyndt F, Glinge C, Postema PG, Amin AS, Nannenberg EA, Ware JS, Whiffin N, Mazzarotto F, Škorić-Milosavljević D, Krijger C, Arbelo E, Babuty D, Barajas-Martinez H, Beckmann BM, Bézieau S, Bos JM, Breckpot J, Campuzano O, Castelletti S, Celen C, Clauss S, Corveleyn A, Crotti L, Dagradi F, de Asmundis C, Denjoy I, Dittmann S, Ellinor PT, Ortuño CG, Giustetto C, Gourraud JB, Hazeki D, Horie M, Ishikawa T, Itoh H, Kaneko Y, Kanters JK, Kimoto H, Kotta MC, Krapels IPC, Kurabayashi M, Lazarte J, Leenhardt A, Loeys BL, Lundin C, Makiyama T, Mansourati J, Martins RP, Mazzanti A, Mörner S, Napolitano C, Ohkubo K, Papadakis M, Rudic B, Molina MS, Sacher F, Sahin H, Sarquella-Brugada G, Sebastiano R, Sharma S, Sheppard MN, Shimamoto K, Shoemaker MB, Stallmeyer B, Steinfurt J, Tanaka Y, Tester DJ, Usuda K, van der Zwaag PA, Van Dooren S, Van Laer L, Winbo A, Winkel BG, Yamagata K, Zumhagen S, Volders PGA, Lubitz SA, Antzelevitch C, Platonov PG, Odening KE, Roden DM, Roberts JD, Skinner JR, Tfelt-Hansen J, van den Berg MP, Olesen MS, Lambiase PD, Borggrefe M, Hayashi K, Rydberg A, Nakajima T, Yoshinaga M, Saenen JB, Kääb S, Brugada P, Robyns T, Giachino DF, Ackerman MJ, Brugada R, Brugada J, Gimeno JR, Hasdemir C, Guicheney P, Priori SG, Schulze-Bahr E, Makita N, Schwartz PJ, Shimizu W, Aiba T, Schott JJ, Redon R, Ohno S, Probst V, Behr ER, Barc J and Bezzina CR. Enhancing rare variant interpretation in inherited arrhythmias through quantitative analysis of consortium disease cohorts and population controls. Genet Med. 2021;23:47–58.

13. Glazer AM, Kroncke BM, Matreyek KA, Yang T, Wada Y, Shields T, Salem JE, Fowler DM and Roden DM. Deep Mutational Scan of an SCN5A Voltage Sensor. Circ Genom Precis Med. 2020;13:e002786.

14. Cheng J, Novati G, Pan J, Bycroft C, Žemgulytė A, Applebaum T, Pritzel A, Wong LH, Zielinski M, Sargeant T, Schneider RG, Senior AW, Jumper J, Hassabis D, Kohli P and Avsec Ž. Accurate proteome-wide missense variant effect prediction with AlphaMissense. Science. 2023;381:eadg7492.

15. Ioannidis NM, Rothstein JH, Pejaver V, Middha S, McDonnell SK, Baheti S, Musolf A, Li Q, Holzinger E, Karyadi D, Cannon-Albright LA, Teerlink CC, Stanford JL, Isaacs WB, Xu J, Cooney KA, Lange EM, Schleutker J, Carpten JD, Powell IJ, Cussenot O, Cancel-Tassin G, Giles GG, MacInnis RJ, Maier C, Hsieh CL, Wiklund F, Catalona WJ, Foulkes WD, Mandal D, Eeles RA, Kote-Jarai Z, Bustamante CD, Schaid DJ, Hastie T, Ostrander EA, Bailey-Wilson JE, Radivojac P, Thibodeau SN, Whittemore AS and Sieh W. REVEL: An Ensemble Method for Predicting the Pathogenicity of Rare Missense Variants. Am J Hum Genet. 2016;99:877–885.

16. Bergquist T, Stenton SL, Nadeau EAW, Byrne AB, Greenblatt MS, Harrison SM, Tavtigian SV, O’Donnell-Luria A, Biesecker LG, Radivojac P, Brenner SE and Pejaver V. Calibration of additional computational tools expands ClinGen recommendation options for variant classification with PP3/BP4 criteria. bioRxiv. 2024:2024.09.17.611902.

17. Starita LM, Ahituv N, Dunham MJ, Kitzman JO, Roth FP, Seelig G, Shendure J and Fowler DM. Variant Interpretation: Functional Assays to the Rescue. Am J Hum Genet. 2017;101:315–325.

18. Glazer AM, Wada Y, Li B, Muhammad A, Kalash OR, O’Neill MJ, Shields T, Hall L, Short L, Blair MA, Kroncke BM, Capra JA and Roden DM. High-Throughput Reclassification of SCN5A Variants. Am J Hum Genet. 2020;107:111–123.

19. Ma JG, O’Neill MJ, Richardson E, Thomson KL, Ingles J, Muhammad A, Solus JF, Davogustto G, Anderson KC, Shoemaker MB, Stergachis AB, Floyd BJ, Dunn K, Parikh VN, Chubb WH, Perrin MJ, Roden DM, Vandenberg JI, Ng CA and Glazer AM. Multisite Validation of a Functional Assay to Adjudicate SCN5A Brugada Syndrome-Associated Variants. Circ Genom Precis Med. 2024;17:e004569.

20. O’Neill MJ, Muhammad A, Li B, Wada Y, Hall L, Solus JF, Short L, Roden DM and Glazer AM. Dominant negative effects of SCN5A missense variants. Genet Med. 2022;24:1238–1248.

21. Brnich SE, Abou Tayoun AN, Couch FJ, Cutting GR, Greenblatt MS, Heinen CD, Kanavy DM, Luo X, McNulty SM, Starita LM, Tavtigian SV, Wright MW, Harrison SM, Biesecker LG and Berg JS. Recommendations for application of the functional evidence PS3/BS3 criterion using the ACMG/AMP sequence variant interpretation framework. Genome Med. 2019;12:3.

22. Karczewski KJ, Francioli LC, Tiao G, Cummings BB, Alföldi J, Wang Q, Collins RL, Laricchia KM, Ganna A, Birnbaum DP, Gauthier LD, Brand H, Solomonson M, Watts NA, Rhodes D, Singer-Berk M, England EM, Seaby EG, Kosmicki JA, Walters RK, Tashman K, Farjoun Y, Banks E, Poterba T, Wang A, Seed C, Whiffin N, Chong JX, Samocha KE, Pierce-Hoffman E, Zappala Z, O’Donnell-Luria AH, Minikel EV, Weisburd B, Lek M, Ware JS, Vittal C, Armean IM, Bergelson L, Cibulskis K, Connolly KM, Covarrubias M, Donnelly S, Ferriera S, Gabriel S, Gentry J, Gupta N, Jeandet T, Kaplan D, Llanwarne C, Munshi R, Novod S, Petrillo N, Roazen D, Ruano-Rubio V, Saltzman A, Schleicher M, Soto J, Tibbetts K, Tolonen C, Wade G, Talkowski ME, Neale BM, Daly MJ and MacArthur DG. The mutational constraint spectrum quantified from variation in 141,456 humans. Nature. 2020;581:434–443.

23. Li Z, Jin X, Wu T, Zhao X, Wang W, Lei J, Pan X and Yan N. Structure of human Na(v)1.5 reveals the fast inactivation-related segments as a mutational hotspot for the long QT syndrome. Proc Natl Acad Sci U S A. 2021;118.

24. Thomson KL, Jiang C, Richardson E, Westphal DS, Burkard T, Wolf CM, Vatta M, Harrison SM, Ingles J, Bezzina CR, Kroncke BM, Vandenberg JI and Ng CA. Clinical interpretation of KCNH2 variants using a robust PS3/BS3 functional patch-clamp assay. HGG Adv. 2024;5:100270.

25. Pejaver V, Byrne AB, Feng BJ, Pagel KA, Mooney SD, Karchin R, O’Donnell-Luria A, Harrison SM, Tavtigian SV, Greenblatt MS, Biesecker LG, Radivojac P and Brenner SE. Calibration of computational tools for missense variant pathogenicity classification and ClinGen recommendations for PP3/BP4 criteria. Am J Hum Genet. 2022;109:2163–2177.

26. Tavtigian SV, Harrison SM, Boucher KM and Biesecker LG. Fitting a naturally scaled point system to the ACMG/AMP variant classification guidelines. Hum Mutat. 2020;41:1734–1737.

27. Jaganathan K, Kyriazopoulou Panagiotopoulou S, McRae JF, Darbandi SF, Knowles D, Li YI, Kosmicki JA, Arbelaez J, Cui W, Schwartz GB, Chow ED, Kanterakis E, Gao H, Kia A, Batzoglou S, Sanders SJ and Farh KK-H. Predicting Splicing from Primary Sequence with Deep Learning. Cell. 2019;176:535–548.e24.

28. McGurk KA, Zhang X, Theotokis P, Thomson K, Harper A, Buchan RJ, Mazaika E, Ormondroyd E, Wright WT, Macaya D, Pua CJ, Funke B, MacArthur DG, Prasad SK, Cook SA, Allouba M, Aguib Y, Yacoub MH, O’Regan DP, Barton PJR, Watkins H, Bottolo L and Ware JS. The penetrance of rare variants in cardiomyopathy-associated genes: A cross-sectional approach to estimating penetrance for secondary findings. Am J Hum Genet. 2023;110:1482–1495.

29. Vutthikraivit W, Rattanawong P, Putthapiban P, Sukhumthammarat W, Vathesatogkit P, Ngarmukos T and Thakkinstian A. Worldwide Prevalence of Brugada Syndrome: A Systematic Review and Meta-Analysis. Acta Cardiol Sin. 2018;34:267–277.

30. O’Neill MJ, Yang T, Laudeman J, Calandranis ME, Harvey ML, Solus JF, Roden DM and Glazer AM. ParSE-seq: a calibrated multiplexed assay to facilitate the clinical classification of putative splice-altering variants. Nat Commun. 2024;15:8320.

31. Kapplinger JD, Tester DJ, Alders M, Benito B, Berthet M, Brugada J, Brugada P, Fressart V, Guerchicoff A, Harris-Kerr C, Kamakura S, Kyndt F, Koopmann TT, Miyamoto Y, Pfeiffer R, Pollevick GD, Probst V, Zumhagen S, Vatta M, Towbin JA, Shimizu W, Schulze-Bahr E, Antzelevitch C, Salisbury BA, Guicheney P, Wilde AA, Brugada R, Schott JJ and Ackerman MJ. An international compendium of mutations in the SCN5A-encoded cardiac sodium channel in patients referred for Brugada syndrome genetic testing. Heart Rhythm. 2010;7:33–46.

32. Ishikawa T, Kimoto H, Mishima H, Yamagata K, Ogata S, Aizawa Y, Hayashi K, Morita H, Nakajima T, Nakano Y, Nagase S, Murakoshi N, Kowase S, Ohkubo K, Aiba T, Morimoto S, Ohno S, Kamakura S, Nogami A, Takagi M, Karakachoff M, Dina C, Schott JJ, Yoshiura KI, Horie M, Shimizu W, Nishimura K, Kusano K and Makita N. Functionally validated SCN5A variants allow interpretation of pathogenicity and prediction of lethal events in Brugada syndrome. Eur Heart J. 2021;42:2854–2863.

33. Yamagata K, Horie M, Aiba T, Ogawa S, Aizawa Y, Ohe T, Yamagishi M, Makita N, Sakurada H, Tanaka T, Shimizu A, Hagiwara N, Kishi R, Nakano Y, Takagi M, Makiyama T, Ohno S, Fukuda K, Watanabe H, Morita H, Hayashi K, Kusano K, Kamakura S, Yasuda S, Ogawa H, Miyamoto Y, Kapplinger JD, Ackerman MJ and Shimizu W. Genotype-Phenotype Correlation of *SCN5A* Mutation for the Clinical and Electrocardiographic Characteristics of Probands With Brugada Syndrome. Circulation. 2017;135:2255–2270.

34. Kroncke BM, Glazer AM, Smith DK, Blume JD and Roden DM. SCN5A (Na(V)1.5) Variant Functional Perturbation and Clinical Presentation: Variants of a Certain Significance. Circ Genom Precis Med. 2018;11:e002095.

35. Whiffin N, Minikel E, Walsh R, O’Donnell-Luria AH, Karczewski K, Ing AY, Barton PJR, Funke B, Cook SA, MacArthur D and Ware JS. Using high-resolution variant frequencies to empower clinical genome interpretation. Genet Med. 2017;19:1151–1158.

36. Harrison SM, Dolinsky JS, Knight Johnson AE, Pesaran T, Azzariti DR, Bale S, Chao EC, Das S, Vincent L and Rehm HL. Clinical laboratories collaborate to resolve differences in variant interpretations submitted to ClinVar. Genet Med. 2017;19:1096–1104.

37. Amendola LM, Muenzen K, Biesecker LG, Bowling KM, Cooper GM, Dorschner MO, Driscoll C, Foreman AKM, Golden-Grant K, Greally JM, Hindorff L, Kanavy D, Jobanputra V, Johnston JJ, Kenny EE, McNulty S, Murali P, Ou J, Powell BC, Rehm HL, Rolf B, Roman TS, Van Ziffle J, Guha S, Abhyankar A, Crosslin D, Venner E, Yuan B, Zouk H and Jarvik GP. Variant Classification Concordance using the ACMG-AMP Variant Interpretation Guidelines across Nine Genomic Implementation Research Studies. Am J Hum Genet. 2020;107:932–941.

38. Glazer AM, Davogustto G, Shaffer CM, Vanoye CG, Desai RR, Farber-Eger EH, Dikilitas O, Shang N, Pacheco JA, Yang T, Muhammad A, Mosley JD, Van Driest SL, Wells QS, Shaffer LL, Kalash OR, Wada Y, Bland HT, Yoneda ZT, Mitchell DW, Kroncke BM, Kullo IJ, Jarvik GP, Gordon AS, Larson EB, Manolio TA, Mirshahi T, Luo JZ, Schaid D, Namjou B, Alsaied T, Singh R, Singhal A, Liu C, Weng C, Hripcsak G, Ralston JD, McNally EM, Chung WK, Carrell DS, Leppig KA, Hakonarson H, Sleiman P, Sohn S, Glessner J, e MN, Denny J, Wei WQ, George AL, Jr., Shoemaker MB and Roden DM. Arrhythmia Variant Associations and Reclassifications in the eMERGE-III Sequencing Study. Circulation. 2022;145:877–891.

39. Forrest IS, Chaudhary K, Vy HMT, Petrazzini BO, Bafna S, Jordan DM, Rocheleau G, Loos RJF, Nadkarni GN, Cho JH and Do R. Population-Based Penetrance of Deleterious Clinical Variants. Jama. 2022;327:350–359.

40. Kroncke BM, Smith DK, Zuo Y, Glazer AM, Roden DM and Blume JD. A Bayesian method to estimate variant-induced disease penetrance. PLoS Genet. 2020;16:e1008862.

41. Smits JP, Koopmann TT, Wilders R, Veldkamp MW, Opthof T, Bhuiyan ZA, Mannens MM, Balser JR, Tan HL, Bezzina CR and Wilde AA. A mutation in the human cardiac sodium channel (E161K) contributes to sick sinus syndrome, conduction disease and Brugada syndrome in two families. J Mol Cell Cardiol. 2005;38:969–81.

42. Rossenbacker T, Carroll SJ, Liu H, Kuipéri C, de Ravel TJ, Devriendt K, Carmeliet P, Kass RS and Heidbüchel H. Novel pore mutation in SCN5A manifests as a spectrum of phenotypes ranging from atrial flutter, conduction disease, and Brugada syndrome to sudden cardiac death. Heart Rhythm. 2004;1:610–5.

43. Grant AO, Carboni MP, Neplioueva V, Starmer CF, Memmi M, Napolitano C and Priori S. Long QT syndrome, Brugada syndrome, and conduction system disease are linked to a single sodium channel mutation. J Clin Invest. 2002;110:1201–9.

44. Bezzina C, Veldkamp MW, van Den Berg MP, Postma AV, Rook MB, Viersma JW, van Langen IM, Tan-Sindhunata G, Bink-Boelkens MT, van Der Hout AH, Mannens MM and Wilde AA. A single Na(+) channel mutation causing both long-QT and Brugada syndromes. Circ Res. 1999;85:1206–13.

45. O’Neill MJ, Sala L, Denjoy I, Wada Y, Kozek K, Crotti L, Dagradi F, Kotta MC, Spazzolini C, Leenhardt A, Salem JE, Kashiwa A, Ohno S, Tao R, Roden DM, Horie M, Extramiana F, Schwartz PJ and Kroncke BM. Continuous Bayesian Variant Interpretation Accounts for Incomplete Penetrance among Mendelian Cardiac Channelopathies. Genet Med. 2022.

46. Cerrone M, Remme CA, Tadros R, Bezzina CR and Delmar M. Beyond the One Gene– One Disease Paradigm. Circulation. 2019;140:595–610.

47. Ishikawa T, Masuda T, Hachiya T, Dina C, Simonet F, Nagata Y, Tanck MWT, Sonehara K, Glinge C, Tadros R, Khongphatthanayothin A, Lu T-P, Higuchi C, Nakajima T, Hayashi K, Aizawa Y, Nakano Y, Nogami A, Morita H, Ohno S, Aiba T, Krijger Juárez C, Mauleekoonphairoj J, Poovorawan Y, Gourraud J-B, Shimizu W, Probst V, Horie M, Wilde AAM, Redon R, Juang J- MJ, Nademanee K, Bezzina CR, Barc J, Tanaka T, Okada Y, Schott J-J and Makita N. Brugada syndrome in Japan and Europe: a genome-wide association study reveals shared genetic architecture and new risk loci. European Heart Journal. 2024;45:2320–2332.

48. Bersell KR, Yang T, Mosley JD, Glazer AM, Hale AT, Kryshtal DO, Kim K, Steimle JD, Brown JD, Salem JE, Campbell CC, Hong CC, Wells QS, Johnson AN, Short L, Blair MA, Behr ER, Petropoulou E, Jamshidi Y, Benson MD, Keyes MJ, Ngo D, Vasan RS, Yang Q, Gerszten RE, Shaffer C, Parikh S, Sheng Q, Kannankeril PJ, Moskowitz IP, York JD, Wang TJ, Knollmann BC and Roden DM. Transcriptional Dysregulation Underlies Both Monogenic Arrhythmia Syndrome and Common Modifiers of Cardiac Repolarization. Circulation. 2023;147:824–840.

49. Walsh R, Mauleekoonphairoj J, Mengarelli I, Bosada FM, Verkerk AO, van Duijvenboden K, Poovorawan Y, Wongcharoen W, Sutjaporn B, Wandee P, Chimparlee N, Chokesuwattanaskul R, Vongpaisarnsin K, Dangkao P, Wu CI, Tadros R, Amin AS, Lieve KVV, Postema PG, Kooyman M, Beekman L, Sahasatas D, Amnueypol M, Krittayaphong R, Prechawat S, Anannab A, Makarawate P, Ngarmukos T, Phusanti K, Veerakul G, Kingsbury Z, Newington T, Maheswari U, Ross MT, Grace A, Lambiase PD, Behr ER, Schott JJ, Redon R, Barc J, Christoffels VM, Wilde AAM, Nademanee K, Bezzina CR and Khongphatthanayothin A. A Rare Noncoding Enhancer Variant in SCN5A Contributes to the High Prevalence of Brugada Syndrome in Thailand. Circulation. 2024.

50. O’Neill MJ, Wada Y, Hall LD, Mitchell DW, Glazer AM and Roden DM. Functional Assays Reclassify Suspected Splice-Altering Variants of Uncertain Significance in Mendelian Channelopathies. Circ Genom Precis Med. 2022;15:e003782.

