## Supplemental Figures for "Cohort-scale automated patch clamp data improves variant classification and penetrance stratification for *SCN5A*-Brugada Syndrome"

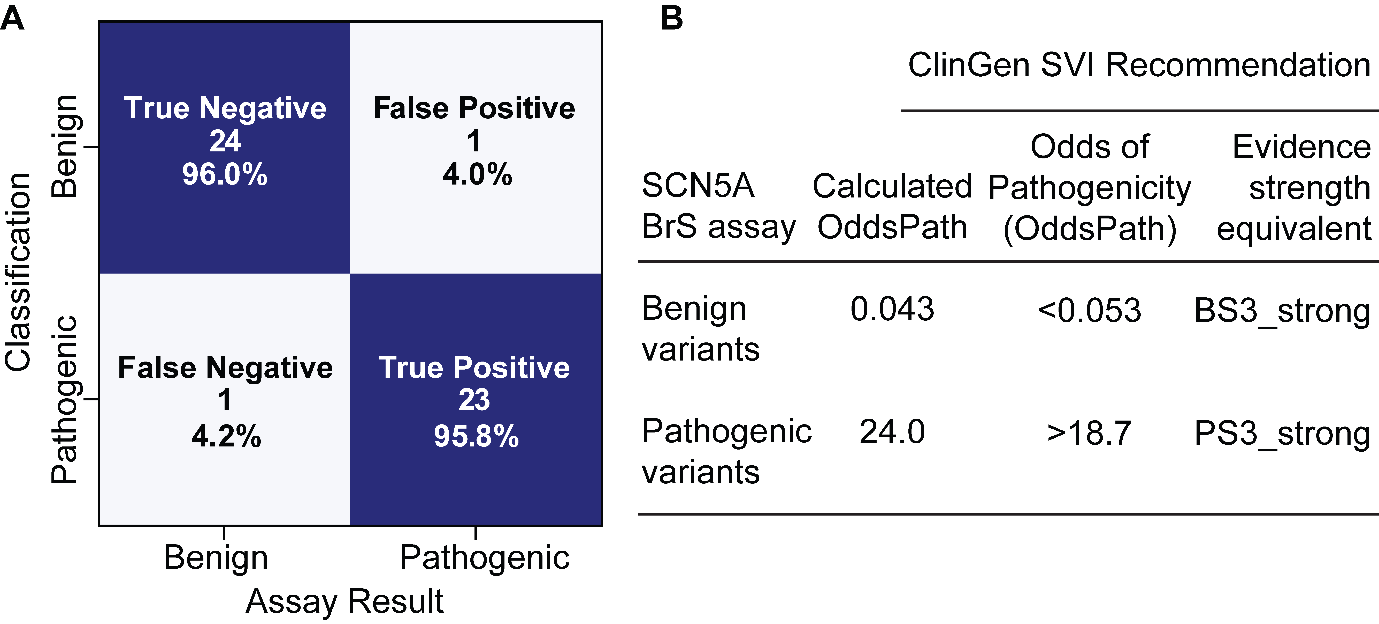

### **Figure S1: Multisite OddsPath Recalibration.**

OddsPath scores post recalibration of SCN5A-BrS physiological voltage assay in accordance with ClinGen SVI Working Group guidelines^1^. This assay can provide evidence at strong levels for both BS3 and PS3, benign and pathogenic, but will be conservatively applied at BS3_moderate in this assay.

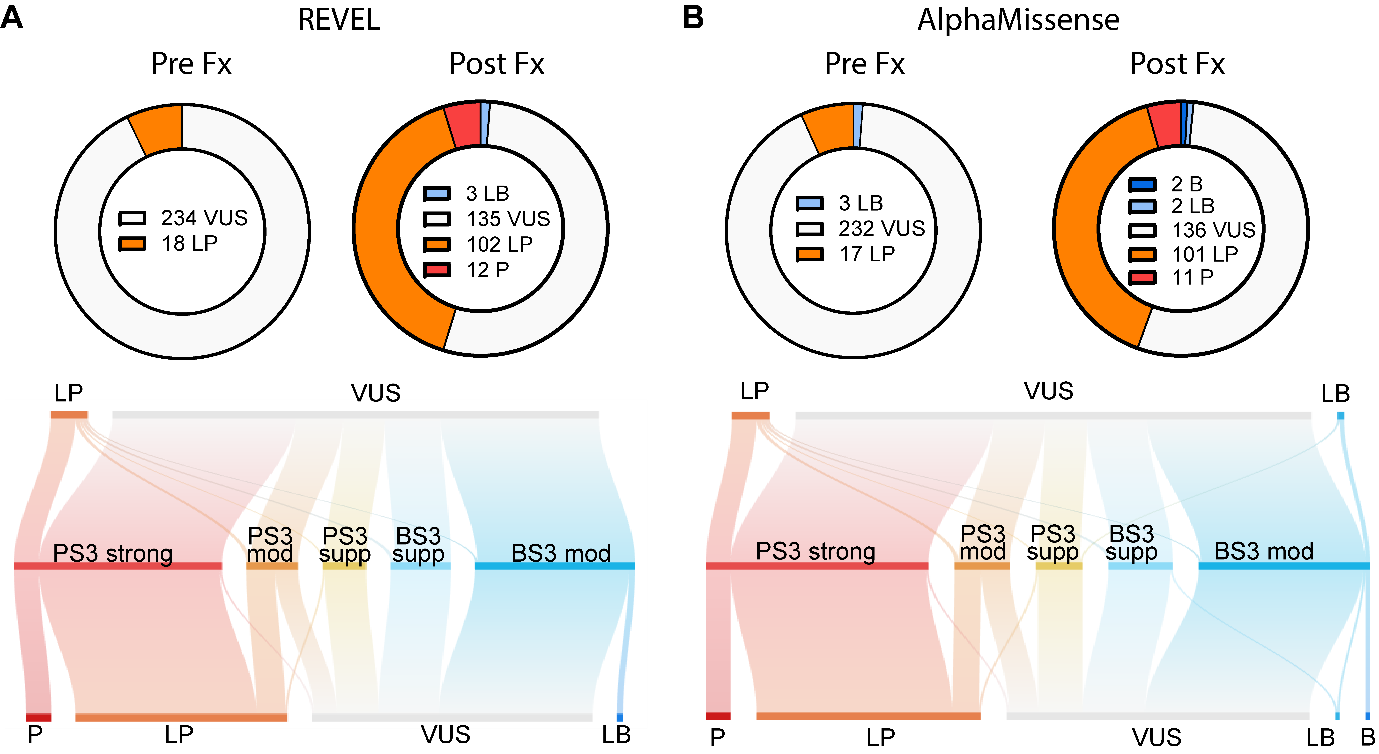

### **Figure S2: Classification of variants with or without functional data.**

Sensitivity analysis show variants in this cohort classified with or without functional data and with alternate *in silico* evidence. In this figure, in silico evidence is applied at standard supporting evidence strengths (PP3)^2^ pre- and post-functional data using REVEL at PP3 (A) and AlphaMissense at PP3. Classifications using calibrated in silico evidences found in Figure 3.

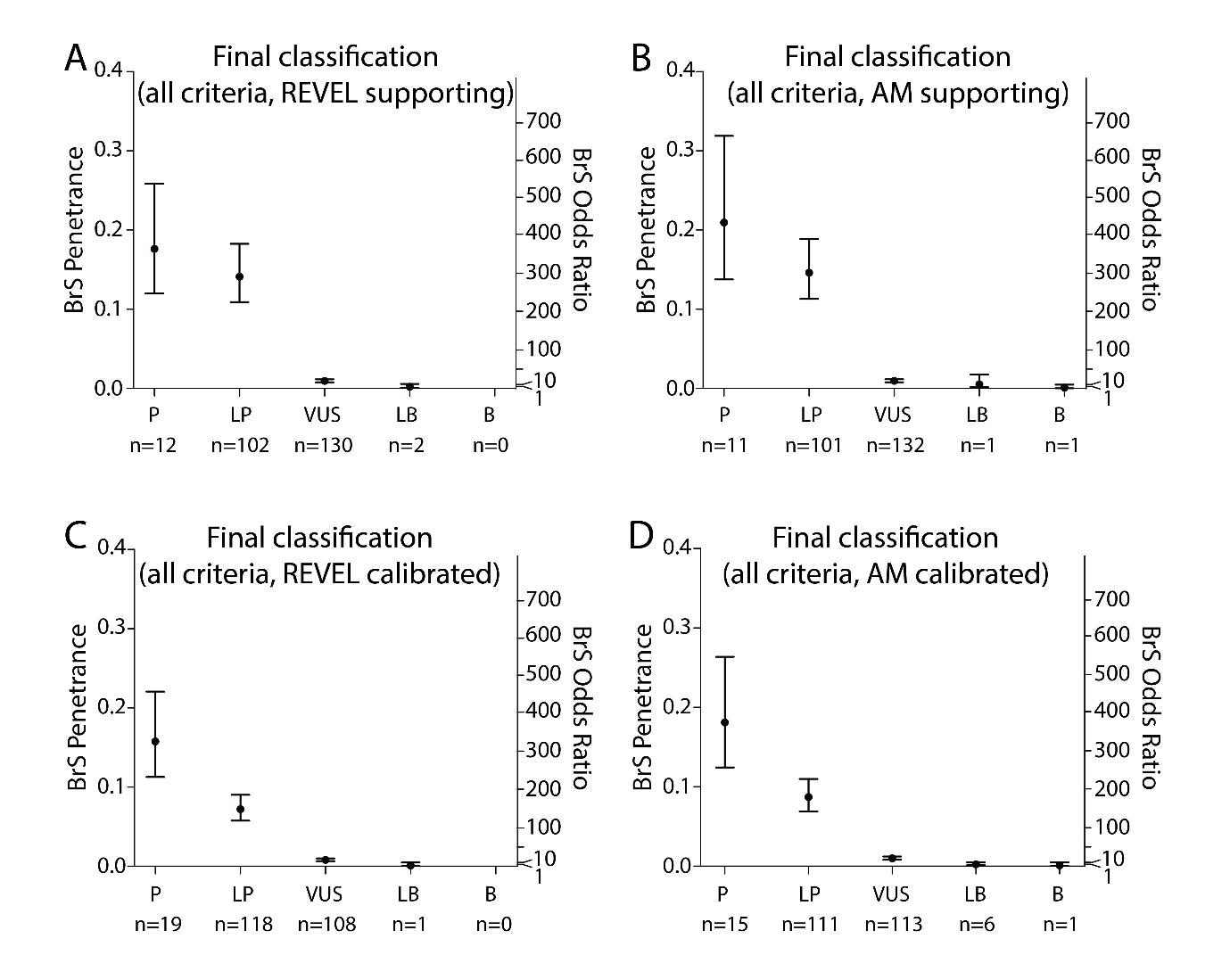

### **Figure S3: Penetrance estimates for final classifications.**

Penetrance estimates for final classifications using REVEL at supporting level (A), AlphaMissense at supporting level (B), calibrated REVEL and calibrated AlphaMissense. See Table S10 for data.

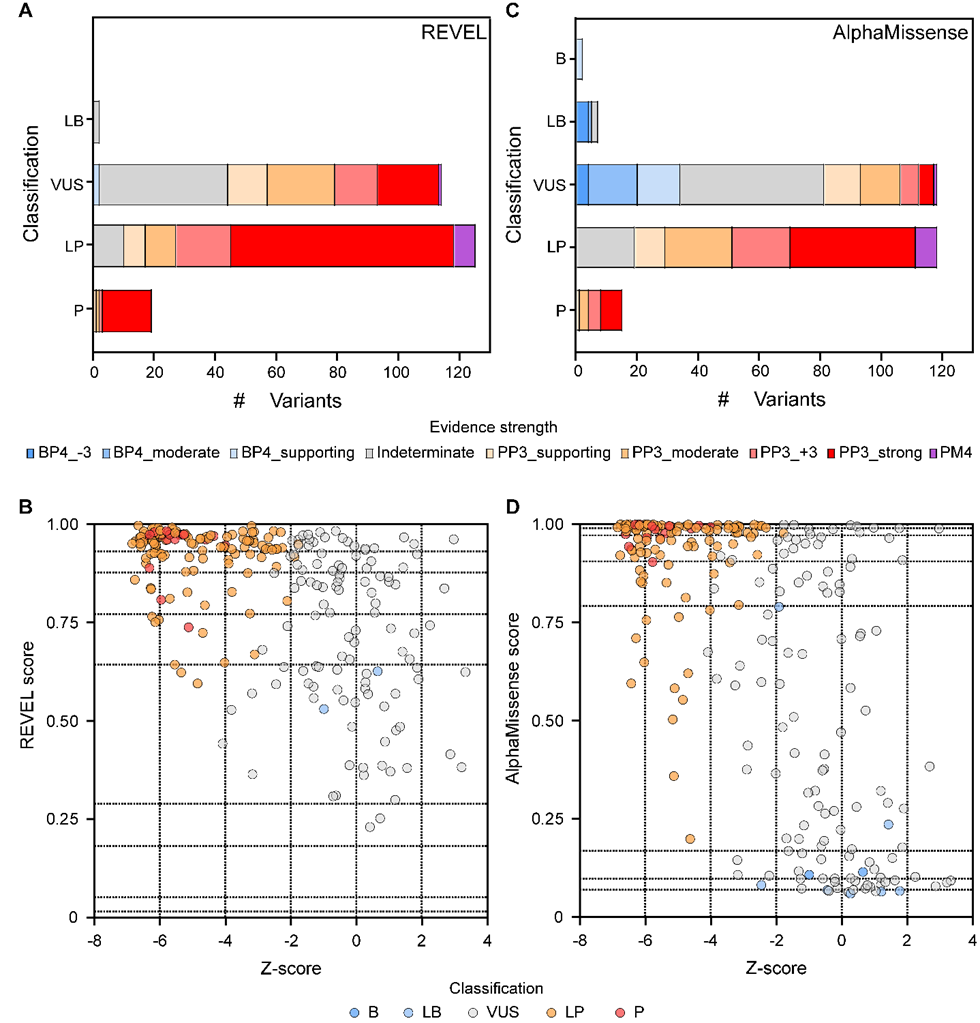

### **Figure S4: Calibrated computational evidence used in this study, segregated by final classification and by *Z*-score.**

The strength of calibrated REVEL (A) and AlphaMissense (C) *in silico* evidence applied per variant in each final classification category. REVEL (B) and AlphaMissense (D) scores plotted against functional evidence Z-score, colored by final classification.

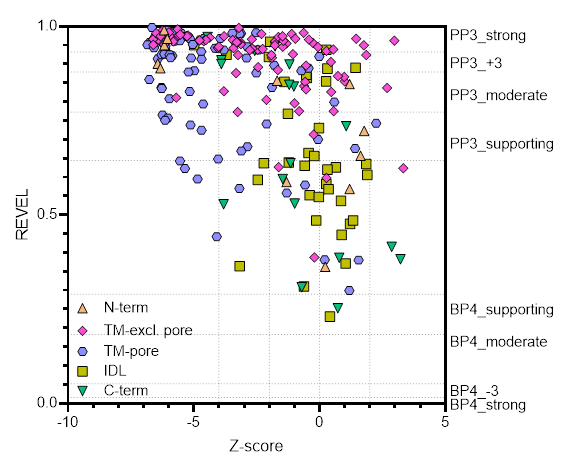

### **Figure S5: Calibrated REVEL computational by *Z*-score, grouped by location in protein.**

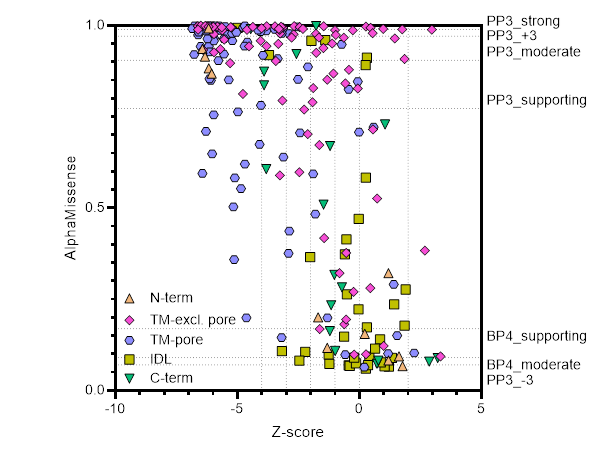

### **Figure S6: Calibrated AlphaMissense computational by *Z*-score, grouped by location in protein.**

### **References**

1	Brnich, S. E. *et al.* Recommendations for application of the functional evidence PS3/BS3 criterion using the ACMG/AMP sequence variant interpretation framework. *Genome Med* **12**, 3, doi:10.1186/s13073-019-0690-2 (2020).

2	Richards, S. *et al.* Standards and guidelines for the interpretation of sequence variants: a joint consensus recommendation of the American College of Medical Genetics and Genomics and the Association for Molecular Pathology. *Genetics in Medicine* **17**, 405-423, doi:10.1038/gim.2015.30 (2015).
